# Precision Estimates of Longitudinal Brain Aging Capture Unexpected Individual Differences in One Year

**DOI:** 10.1101/2025.02.21.25322553

**Authors:** Maxwell L. Elliott, Jingnan Du, Jared A. Nielsen, Lindsay C. Hanford, Pia Kivisäkk, Steven E. Arnold, Bradford C. Dickerson, Ross W. Mair, Mark C. Eldaief, Randy L. Buckner

**Author notes:** Correspondence (M.L.E), (R.L.B).

## Abstract

Longitudinal studies are required to measure individual differences in human brain aging, but they are dif>icult to estimate over short intervals because of measurement error. Using cluster scanning, an approach that reduces error by densely repeating rapid structural scans, we assessed brain aging in individuals across three longitudinal timepoints spaced across one year. Cluster scanning substantially improved the precision of individualized estimates, revealing previously undetectable individual differences in brain change. In just one year, expected differences in the rates of brain aging between younger and older individuals were evident, as were differences between cognitively unimpaired and impaired individuals. Each person’s brain change trajectory was compared to modeled normative expectations from a large cohort of age-matched UK Biobank participants. Cognitively unimpaired older individuals variably revealed relative brain maintenance, unexpectedly rapid decline, and asymmetrical changes. These atypical brain aging trajectories were found across structures and veri>ied in independent within-individual test and retest data. Cluster scanning promises to advance our understanding of the marked heterogeneity in brain aging by affording better short-term tracking of individual variability in structural change.

## Introduction

There are large individual differences in the rate of brain aging in older adults, even among individuals of the same age and neurodegenerative disease diagnosis (e.g., Arnold et al., 2024, Cook et al., 2017; Ferreira et al., 2020; Ossenkoppele et al., 2016; Nyberg et al., 2012; 2023; Raz et al., 2005). Faster age-related brain changes in older adults are linked to cognitive decline and begin years before dementia onset in individuals with neurodegenerative diseases (Cox et al., 2021; Jack et al., 2018; Nyberg et al., 2023). Longitudinal studies are necessary to assess these brain changes in individuals (Lindenberger et al., 2011; Oschwald et al., 2020; Schaie 1967). However, standard longitudinal MRI estimates fail to track individual trajectories of brain change over short time intervals because the measurement error is often larger than the typical amount of change. For example, standard estimates of hippocampal volume – one of the most commonly used markers of brain aging – have a measurement error of approximately 2-5% (Tustison et al., 2014; Vidal-Pineiro et al., 2024), while the average annual rate of hippocampal change is ∼1-3% in cognitively unimpaired older adults and ∼3-5% in older adults with probable Alzheimer’s disease (AD; Barnes et al., 2009; Jack et al., 2000; 2008; Pini et al., 2016). Therefore, in a yearlong study of hippocampal atrophy, the amount of measurement error will typically be as large or larger than the expected amount of true change, making the measurements unsuitable for confidently characterizing brain aging within individuals.

The challenge of measurement error in the study of brain aging has been addressed in two ways. First, longitudinal measurements from many individuals are averaged or modeled together to stabilize estimates. This approach is frequently used to estimate normative aging trajectories, examine group differences, and study the average impact of interventions (e.g., Barnes et al., 2009; Bethlehem et al., 2022; Jack et al., 2000; Sabuncu et al., 2011; Storsve et al., 2014; van Dyck et al., 2022). Another approach is to track individuals over long intervals so that the size of the expected change is larger than the error (e.g., Driscoll et al., 2009; Fujita et al., 2023; Nyberg et al., 2020). However, extended longitudinal studies are slow and expensive.

A third untested approach, explored for the first time in this paper, seeks to reduce within-individual measurement error by densely repeating brain scans at each longitudinal timepoint. Every morphometric estimate is a combination of the underlying structure size and measurement error (Crocker & Algina, 1986; Kuder & Richardson, 1937). When just a single scan is acquired at each timepoint, measurement error is necessarily dealt with at the group level or across timepoints. An alternative approach is to collect repeated structural measurements at each timepoint to directly sample measurement variability and reduce error through within-individual averaging or modeling. High-precision estimates of brain aging could facilitate research and translational endeavors including more efficient study designs, individualized assessments, within-individual treatment response monitoring, and faster futility signals in clinical trials. A barrier to collecting dense, repeat samples within individuals is the burden of long MRI acquisition protocols.

Relevant to this goal, advances in MRI acceleration now allow high-resolution structural MRI scans to be acquired in as little as one minute in contrast to the conventional 5-8 minutes (Bilgic et al., 2015; Dieckmeyer et al., 2021; Mussard et al., 2020; Polak et al., 2018). We previously demonstrated that rapid scans have comparable morphometric measurement error to longer standard scans (Elliott et al., 2023) and further that pooling estimates from multiple rapid scans (“cluster scanning”) can substantially reduce measurement error (Nielsen et al., 2019; Elliott et al., 2024). This design is similar to the “burst sampling” approaches applied in cognitive and behavioral settings (e.g., Nesselroade, 1991; Shiffman et al., 2008; Sliwinski, 2008). With cluster scanning, it may be possible to confidently estimate the rate of brain aging in individuals over brief intervals.

Here we conducted a yearlong longitudinal study of brain aging using cluster scanning (N=38). We assessed individual differences in the rate of brain aging in young to middle-aged adults (labeled Adults) and separately in a diverse group of older adults, including cognitive unimpaired (CU) individuals, individuals with amnestic mild cognitive impairment (MCI), probable Alzheimer’s Disease (AD), and dementia due to frontotemporal lobar degeneration (FTLD). We collected both standard and multiple rapid structural MRI scans across three longitudinal timepoints (48 rapid scans per individual). Furthermore, test and retest MRI sessions on separate days were collected at each timepoint to directly assess the amount of error in the longitudinal measurements of brain change. We benchmarked these high-precision individualized estimates to normative expectations of brain aging derived from the longitudinal UK Biobank sample (N=4,243). We discovered that stable individual-specific estimates of brain change can be detected within one year, revealing both expected differences between older and younger adults and those with cognitive impairment. Of most interest, we found considerable, unexpected variation in brain aging among cognitively unimpaired individuals.

## Results

### Normative brain aging trajectories

Cross-sectional differences and longitudinal changes in brain aging were estimated from the large UK Biobank sample (Figure 1). We initially focused on the hippocampus given its role in memory and its non-linear aging trajectory (i.e., minimal atrophy in younger adults followed by greater atrophy in advanced aging; Fjell et al., 2013; Raz et al., 2004). The data are plotted in five-year age bins to emphasize the age-related differences and to highlight the variability around the means. Plots of brain structures beyond the hippocampus are available in Supplemental Fig. S1.

**Figure 1.**
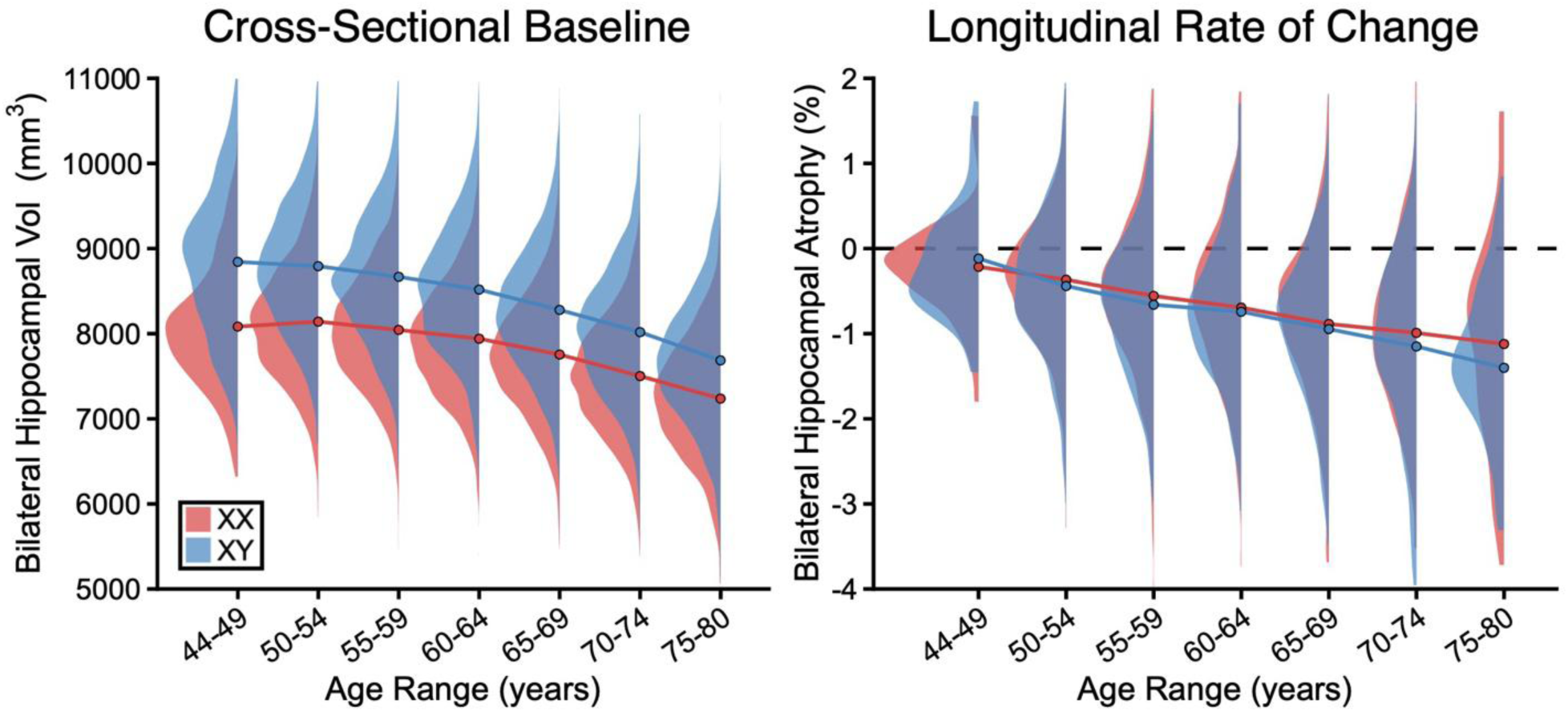
Normative expectations of brain aging. Cross-sectional baseline hippocampal volumes **(Left)** and annualized longitudinal rates of hippocampal change **(Right)** are illustrated for data from the UK Biobank as an example of brain aging. The means and distributions are split by 5-year age bins and plotted separately for genetic males (XY) and females (XX). Mean estimates for each age bin are marked by points connected by lines, while the group distributions are illustrated by density curves. Age-related change in the hippocampus accelerates with advancing age for both sexes. For cross-sectional estimates the sexes differ in proportion to head size. Sex differences are not present in the longitudinal estimates which reference each individual to her or his own head size. The tremendous variability in the estimates at each age bin includes true differences in brain structure as well as measurement error. These normative estimates, which are available for every brain structure, serve as normative (reference) expectations to which individuals can be compared.

Mean hippocampal volume is smaller in older as compared to younger individuals for both sexes. There is a shift between sexes reflecting the established relation between head size and local regional volumes. By contrast, the longitudinal rates of change, which reflect normalized within-individual brain aging trajectories, do not reveal a sex difference but do show the robust acceleration expected with advancing age. Middle-aged individuals largely maintain their hippocampal volume on average (-0.2% per year change in 44-49 year-olds), while older participants show marked atrophy (-1.3% per year change in 75-80 year-olds). These normative data provide estimates of the expected longitudinal rate of change given a person’s chronological age and will be used in upcoming analyses as a reference for precision measurements made within individuals.

The other notable feature of these data is the tremendous dispersion of estimates around the age-binned means. This dispersion is a combination of measurement error and true between-participant differences in brain aging. Specifically, the dispersion within each age bin is far greater than the expected change over a few years, illustrating why it is difficult to track brain aging in individuals over short longitudinal intervals. Our upcoming analyses adopt a cluster scanning approach to increase the precision of within-individual estimates.

### Within-individual change can be reliably detected within one year using cluster scanning

Standard MRI structural scans, as illustrated above, are useful for determining the rate of brain aging when estimates are averaged over people (Figure 1) or when estimated from extended longitudinal epochs. Over short intervals, standard estimates are not useful at the individual level because the amount of measurement error is typically larger than the amount of change. To enable within-individual precision measurement, we conducted a yearlong longitudinal study in which eight rapid structural scans (1’12’’ each), accelerated with compressed sensing, were collected at each of six study visits for a total of 48 scans per person (Figure 2). These visits were temporally staggered to separate the estimate of the yearlong longitudinal change from the estimate of the test-retest measurement error. We hypothesized that individualized trajectories of brain aging could be assessed within a year by pooling measurements from multiple rapid structural MRI scans.

**Figure 2.**
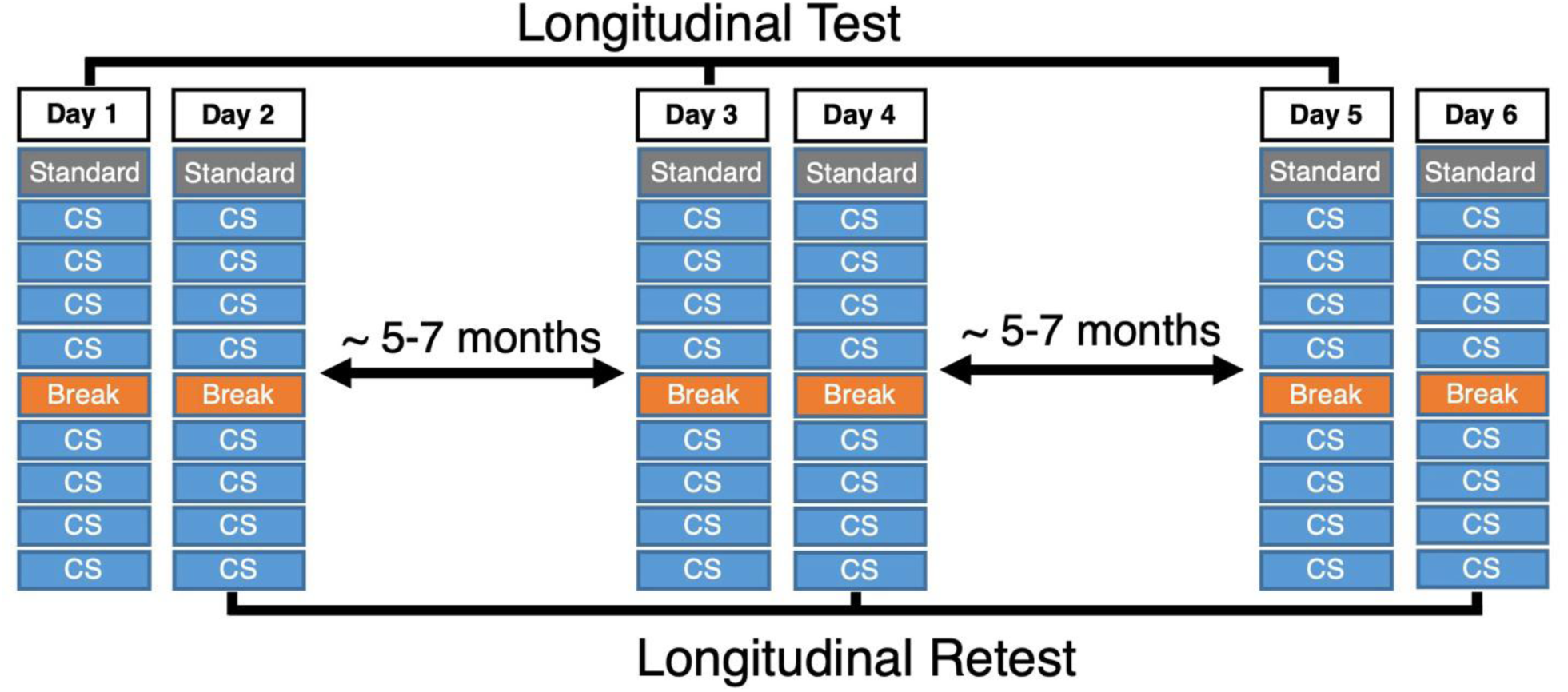
Study design to estimate precision brain aging fully within individuals. The study design combined test- retest sessions at 3 longitudinal timepoints with repeat MRI acquisitions to (1) stabilize within-individual brain morphometric measures and (2) allow for direct estimation of measurement error. In each scanning session, a single standard scan (acquisition time = 5’12’’; gray boxes) and 8 rapid scans (acquisition time = 1’12’’ per scan; blue boxes) were collected. Each scan session contained a short break (orange box) halfway through to reduce participant burden and reposition the head. Each participant was scanned on 6 separate days with test-retest pairs of sessions occurring close in time at spaced longitudinal timepoints. Having test-retest sessions at each longitudinal timepoint allowed for independent measurements of change for each participant (labeled “Longitudinal Test” and “Longitudinal Retest”).

We first explored test-retest measurement error (extending Elliott et al., 2024). The mean measurement error for hippocampal volume from a single rapid structural scan (left hippocampus = 92.4 mm^3^ or 3.4%, right hippocampus = 82.9 mm^3^ or 2.3%) was nearly the same as the amount of error from a standard longer (5’12’’) structural scan (left hippocampus = 99.1 mm^3^ or 3.4%, right hippocampus = 80.8 mm^3^ or 2.2%). Pooling estimates across 2, 4, and 8 rapid scans reduced test-retest measurement error. On average, the measurement error from eight rapid scans (left hippocampus = 33.2 mm^3^ or 1.0%, right hippocampus = 39.0 mm^3^ or 1.1%) was nearly 3 times smaller (Figure 3a left) than a standard scan. Critically, this pattern was mirrored for longitudinal estimates of the rate of hippocampal atrophy across a year (Figure 3a right) and was found across numerous other measures of volume and thickness (see Supplemental Fig. S2).

**Figure 3.**
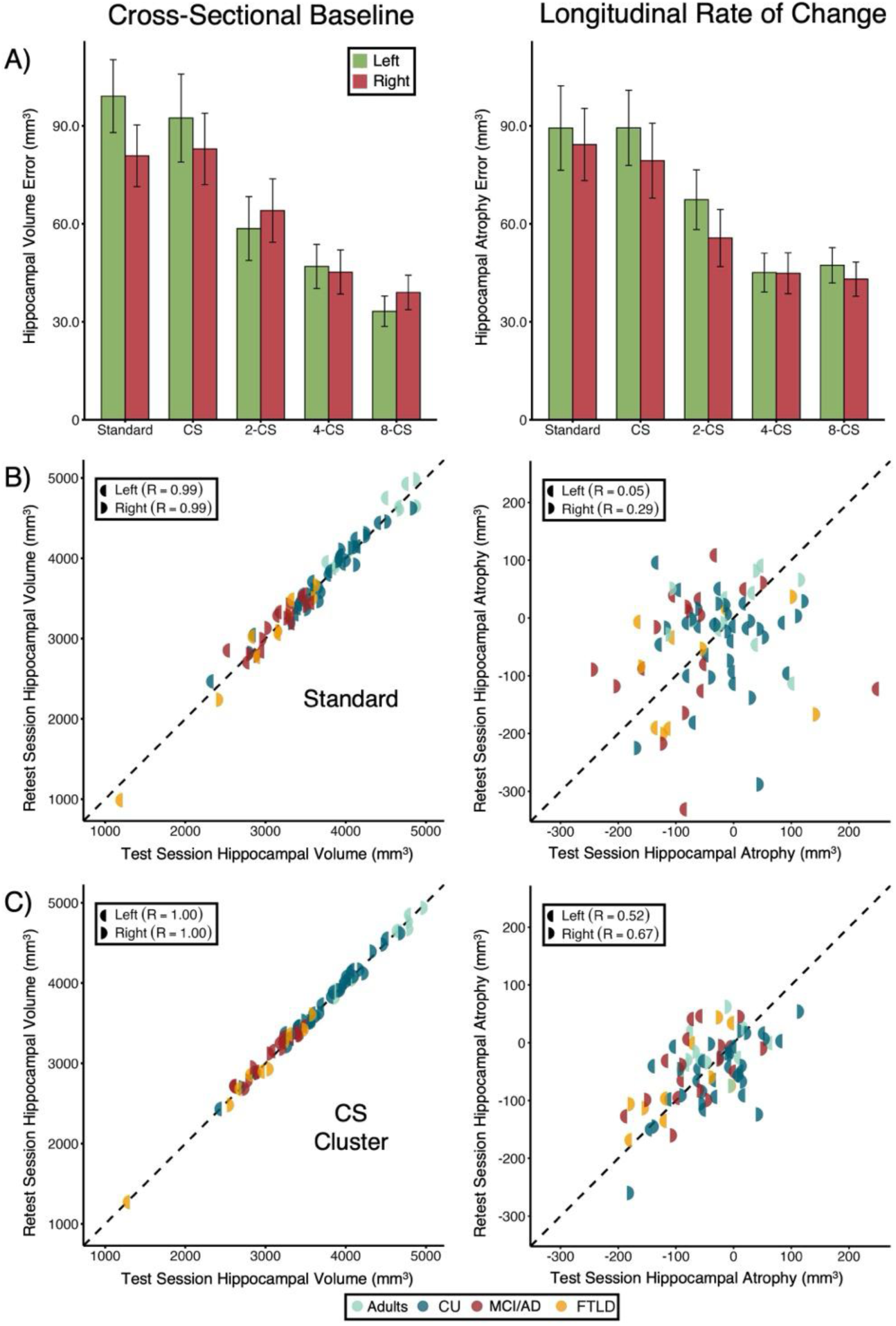
Cluster scanning enables stable estimates of brain aging in one year. **(A)** Absolute cross-sectional measurement error of hippocampal volume **(Left)** and longitudinal measurement error of hippocampal rate of change **(Right)** are illustrated for a single standard T_1_-weighted structural scan as compared to a single (labeled CS) or multiple (labeled 2-CS, 4-CS, and 8-CS) rapid structural scans. Error bars represent the standard error of the mean. Note how measurement error decreases when multiple rapid scans are pooled for both cross-sectional and longitudinal estimates. **(B)** The individual participant values are plotted for cross-sectional test and retest hippocampal volumes **(Left)** and longitudinal test and retest hippocampal change rates **(Right)** for standard scans. For the cross-sectional estimates, the dispersion re]lects the large baseline differences between participants’ hippocampal volumes. The high correlation does not translate into stable estimates of longitudinal change, which are an order of magnitude smaller than the between-individual differences. **(C)** Pooled estimates from 8 rapid scans, by contrast, show moderate longitudinal reliability and clear structure indicating the estimates are capturing true between-individual variability in the longitudinal rate of change over a year. R values are the Pearson’s correlation coef]icient. Colors demarcate the participant groups (Adults; CU = Cognitively Unimpaired older adults; MCI = Mild Cognitive Impairment, AD = probable Alzheimer’s Disease; FTLD = Frontotemporal Lobar Degeneration). Left half circles are estimates from the left hemisphere and right half circles are from the right hemisphere.

Estimates of measurement reliability were explored next. The cross-sectional test-retest reliability of hippocampal volume was high for both the standard structural scan and the pooled estimates (r>0.99; Figure 3b left). This high level of cross-sectional test-retest reliability reflects the stability of estimates in the context of large between-individual baseline differences in hippocampal volume (range from ∼1000 to 5000 mm^3^). While the cross-sectional reliability is better with the pooled rapid scan estimates (Figure 3c left), the benefit is hard to appreciate in the context of between-individual variance. The difference between the two approaches becomes clear when longitudinal change is explored, where the amount of change in a year within most individuals is considerably smaller than baseline differences between people.

The test-retest reliability of longitudinal differences in hippocampal atrophy with standard scanning was poor (left r = 0.05, right r = 0.29; Figure 3b right). It is difficult to see any reliable structure in the longitudinal estimates, consistent with the field’s consensus that, in the absence of rapid neurodegenerative decline, change within individuals is not detectable over short intervals because of measurement error. By contrast, clear structure emerged when pooled estimates were examined (left r = 0.52, right r = 0.67; Figure 3c right) with the hippocampal longitudinal rates of change ranging from ∼100 mm^3^ of growth to 300 mm^3^ of atrophy. The benefits from cluster scanning exemplified by hippocampal volume were also apparent across many other measures of interest to brain aging (Supplemental Figs. S3-S6).

Thus, by driving down measurement error with cluster scanning, we could reliably assess individual differences in brain aging in one year that were not apparent using standard MRI protocols. In upcoming analyses, we used all available rapid scans to stabilize the within-individual estimates of longitudinal change.

### Precision estimates of hippocampal atrophy reveal expected differences

To detect individualized trajectories of brain aging, we estimated the rate of change using all 48 rapid scans from all study visits (note that this is approximately one hour of total scan time). We first evaluated the sensitivity of cluster scanning by comparing the trajectories of participants from the four groups in our study (adults, cognitively unimpaired older adults, individuals with amnestic MCI and AD, and FTLD), each with a different expected trajectory of change in brain structure.

The measured trajectories broadly followed expectations based on their cognitive status at baseline (Figure 4). Plots from individuals illustrate that the estimates are stable across independent test-retest data acquired on separate days (Figure 5). Adults below the age of 50 had little hippocampal atrophy across the year (median annualized change = -0.3%, range -1.6% to 1.4%), near the expected normative mean rate of - 0.2% (SD = 0.6%) for the youngest group (44-49 year-olds; Figure 1). This expected slower rate of hippocampal atrophy in younger adults was exemplified by the 25-29 year-old male in our study who had -0.1% (95% CI = -0.9 to 0.6) and -0.5% (95% CI = -1.2 to 0.2) annualized rates of atrophy in the left and right hippocampus, respectively (Figures 4a and 5a). Younger individuals in our sample showed minimal hippocampal atrophy.

**Figure 4.**
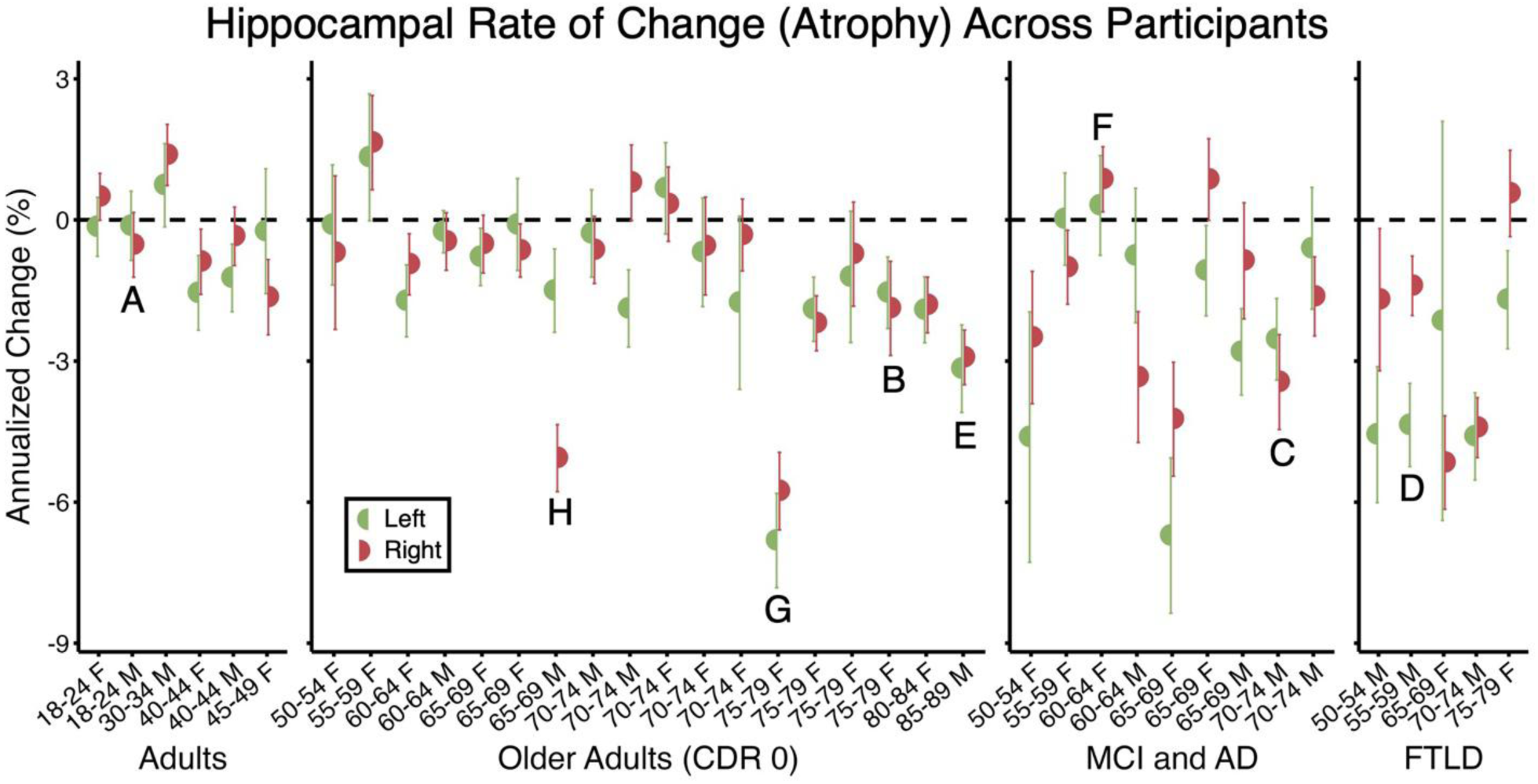
Precision estimates of longitudinal hippocampal change from one year are highly variable across people. Estimates of the annualized rate of left (green half circles) and right (red half circles) hippocampal volume rate of change over a year for every participant in the longitudinal cluster scanning study. The high precision rate of change estimates was derived from all 48 rapid structural scans for each participant. Error bars represent 95% con]idence intervals. Participants are grouped by age and clinical diagnosis and labeled by age and sex: Adults; Cognitively Unimpaired Older Adults; Mild Cognitive Impairment (MCI) / probable Alzheimer’s Disease (AD); Frontotemporal Lobar Degeneration (FTLD). CDR = Clinical Dementia Rating; M = male; F = female. Note the differences between groups and the tremendous variability within groups, including within the Cognitively Unimpaired Older Adults. Letter labels refer to speci]ic participants plotted in more detail in Figure 5.

**Figure 5.**
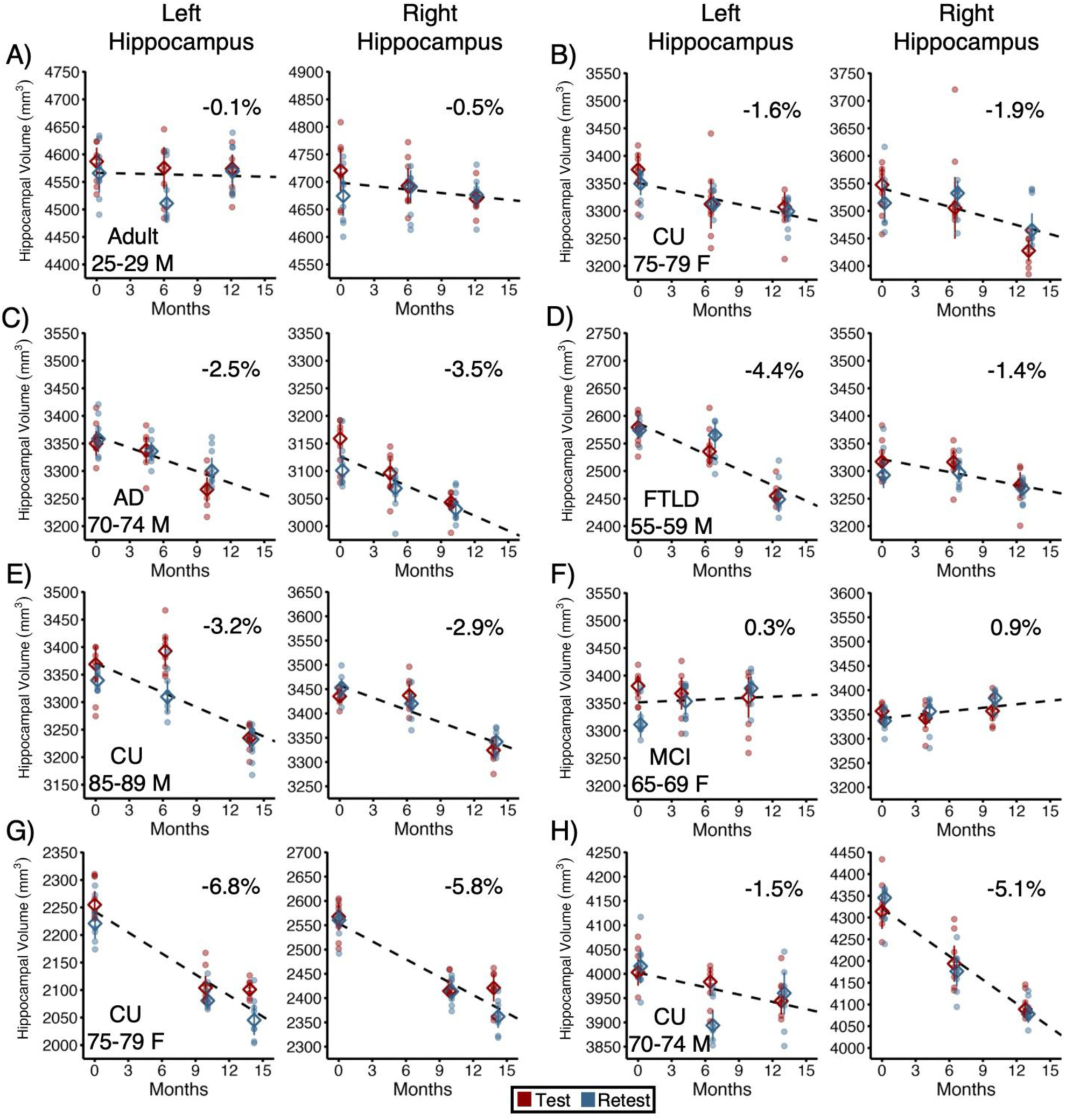
Case studies reveal variability between individuals in the rate and pattern of hippocampal change. Longitudinal rate of change estimates are plotted in detail for eight participants (**A-H**) to illustrate variability between participants. A to H correspond to the participants labeled in Figure 4. For each participant, the pair of panels separately plot the left and right hippocampal volumes, with every individual scan estimate displayed from all 48 scans acquired across the 6 separate days of scanning. Within each test-retest session pair, the ]irst day’s estimates are colored in red and the second day’s estimates are in blue to visualize reliability. Filled circles display the individual scan estimates; open diamonds display the median value for that day with an error bar representing the 95% con]idence interval. The estimated annualized rate of change is in the top right corner of each plot. CU = Cognitively Unimpaired Older Adults; MCI = Mild Cognitive Impairment; AD = probable Alzheimer’s Disease; FTLD = Frontotemporal Lobar Degeneration; M = male; F = female.

At the other end of the spectrum, we observed clear and dramatic within-individual rates of atrophy in individuals presenting with cognitive impairment at baseline. We observed hippocampal atrophy above 2.5% in 5 out of 9 individuals diagnosed with MCI or AD (median annualized change = -1.4%, range = -6.7% to 0.9%) and 4 out of 5 individuals diagnosed with FTLD (median annualized change = -3.3%, range = -5.2% to 0.6%). Furthermore, atrophy tended to be roughly symmetric in MCI and AD cases and asymmetric in the FTLD cases, consistent with known progression patterns (Figure 4). Representative examples include a 70-74 year-old male diagnosed with AD who had -2.5% (95% CI = -3.4 to -1.7) and -3.5% (95% CI = -4.5 to -2.4) annualized rates of atrophy in the left and right hippocampus, respectively (Figures 4c and 5c), and the 55-59 year old male with FTLD who had -4.4% (95% CI = -5.2 to -3.5) and -1.4% (95% CI = -2.0 to -0.8) annualized rates of atrophy in the left and right hippocampus, respectively.

Stages of disease progression were evident in individuals with FTLD. For example, in the 65-69 year-old female and the 70-74 year-old male with FTLD, the current rates of atrophy are more symmetric (Figure 4 right). However, in both individuals, the left hippocampus is highly atrophied at baseline, indicative of an advanced disease state where the leading edge of atrophy progresses to the right hemisphere (see Supplemental Figs. S7 and S8). That is, little left hippocampal volume remains to support further atrophy.

Of most interest, we observed substantial heterogeneity in the rates of hippocampal atrophy in cognitively unimpaired older adults (median annualized change = -0.8%, range = -6.8% to 1.6%). Within this heterogeneity, 15 out of 18 participants had left and right hippocampal atrophy rates below 2.5% annualized, which is typical of age-matched rates from the UK Biobank reference sample (Figure 1; e.g., 70–74 year-old UK Biobank group mean rate = 1.1%, SD = 1.1%). A moderate rate of hippocampal atrophy, typical of cognitively unimpaired older adults, is exemplified by a 75-79 year-old female who displayed -1.6% (95% CI = -2.3 to - 0.8) and -1.9% (95% CI = -2.9 to -0.9) annualized rates of atrophy in the left and right hippocampus, respectively (Figures 4b, 5b and 6).

**Figure 6.**
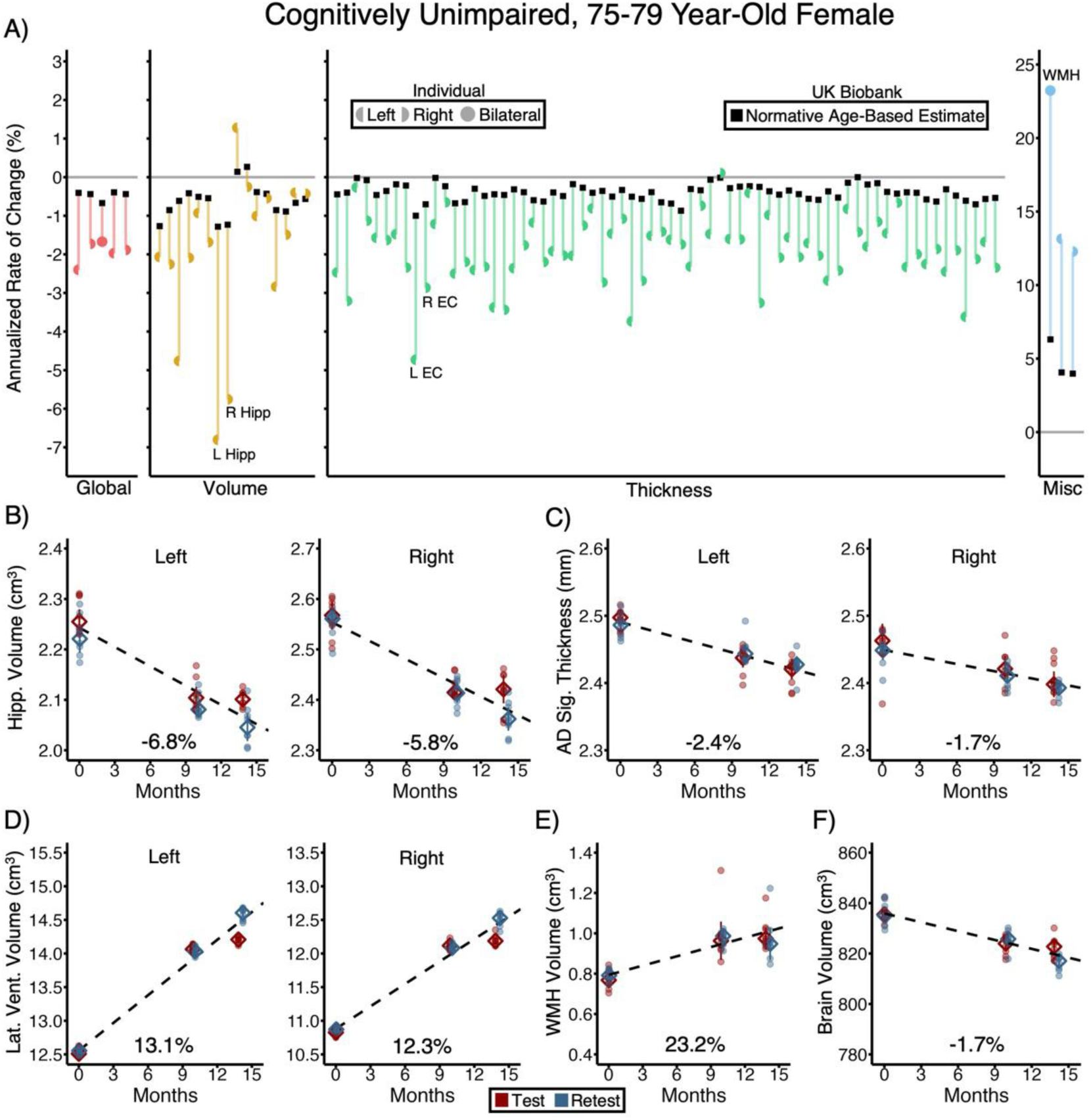
A case study of unexpected brain decline in a cognitively unimpaired 75-79 year-old female. A dashboard visualization of all available brain measures for a single individual is plotted and referenced to normative expectations for similarly aged individuals from the UK Biobank. (**A**) The annualized rate of change for every measure, grouped by type of measure, is displayed in color for the individual (modeled from her 48 scan estimates) in relation to the normative expectation of change for that speci]ic measure labeled by a black square. Each reference value is derived from the 300 UK Biobank participants closest in age to this participant. All reference UK Biobank participants were within 2.7 years of this individual. The colored lines link the individual’s estimates to the normative expectations to reveal how much she deviates. Red = global measures including whole brain volume, whole hemisphere cortical thickness, and cortical thickness in the AD signature regions (see text); Yellow = subcortical volume measures; Green = regional cortical thickness estimates; Blue = white matter hypointensity (WMH) volume and ventricle volumes. Left half circles are estimates from the left hemisphere, right half circles are from the right hemisphere, and whole circles re]lect bilateral measurements (e.g., WMH). See Supplemental Fig. S11 for a full reference of labels for all the morphometrics. This 75-79 year-old female shows accelerated atrophy broadly across the brain, with particularly prominent decline of the hippocampus (Hipp.) and entorhinal cortex (EC). (**Bottom Panels**) Plots show more detail for selected brain morphometric measures using the format of Figure 5 including left and right hippocampal (Hipp.) volumes (**B**), left and right thickness for Alzheimer’s Disease signature (AD Sig.) regions (**C**), left and right lateral (Lat.) ventricle (Vent.) volumes (**D**), WMH (**E**), and global brain volume (**F**). Each panel plots all 48 individual scan estimates. Within each test-retest session pair, the ]irst day’s estimates are colored in red and the second day’s estimates are in blue. Filled circles display the individual scan estimates; open diamonds display the median value for that day with an error bar representing the 95% con]idence interval. Over the year of observation, this participant was diagnosed with cancer and underwent invasive treatment during the gap between her ]irst and second sets of scans. Furthermore, based on follow-up evaluation, this participant was diagnosed with mild cognitive impairment the year following this study. See Supplemental Fig. S12A for a parallel visualization that includes 95% con]idence intervals around each estimate of change.

When considered together, the adult and older adult groups displayed an expected trend of gradually accelerating hippocampal atrophy from the teenagers (Figures 4a and 5a) to the oldest octogenarians (Figures 4b, 4e, 5b, and 5e). However, there were interesting exceptions.

### Unexpected idiosyncratic trajectories of brain aging

We investigated in greater depth several individuals who deviated from expected trajectories. These are not representative cases. One dramatic brain aging trajectory occurred for a 75-79 year-old cognitively unimpaired female with a clinical dementia rating (CDR) of 0 at baseline who had -6.8% (95% CI = -7.8 to -5.8) and -5.8% (95% CI = -6.6 to -4.9) annualized atrophy in the left and right hippocampus, respectively. These were the highest rates of hippocampal atrophy in our sample and came from an individual deemed unimpaired at enrollment (Figures 5g and 6). Further investigation across volume and cortical thickness measures revealed that this decline was not isolated to the hippocampus but reflected a global and robust pattern of accelerated decline (Figure 6). The pattern was especially stark when compared to a similarly aged cognitively unimpaired 75-79 year-old female case study (Figure 5b). Beyond volume estimates of accelerated atrophy, there was also rapid growth in the left (13.1%, 95% CI = 12.4 to 13.8) and right lateral ventricles (12.2%; 95% CI = 11.5 to 13.0) and rapidly increasing white matter hypointensities (23.2%, 95% CI = 16.8 to 29.6).

Consistent with these brain changes, during her study participation, this participant experienced significant medical illness. After completing the baseline MRI scans, she underwent treatment for cancer. Furthermore, after completing the MRI scans, a follow-up assessment conducted by evaluators blind to our brain imaging results noted significant declines in cognition and activities of daily living, and she displayed a decline in objective neuropsychological testing, which led to a diagnosis of amnestic MCI (global CDR = 0.5). Of the 18 cognitively unimpaired older adults, 17 had follow-up clinical evaluations after their baseline cluster MRI scans. She was the only participant who developed amnestic MCI or dementia.

Another clear example of deviation from diagnostic expectations, but in the opposite direction, is the 65- 69 year-old female who unexpectedly maintained both left (0.3%, 95% CI = -0.8 to 1.4) and right (0.9%, 95% CI = 0.2. to 1.6) hippocampal volume during our study despite a baseline diagnosis of amnestic multi-domain MCI that was initially suspected to be due to AD (Figure 5f). Bolstering confidence in the estimates, brain in the left (-4.3%, 95% CI = -5.2 to -3.5) and right (-4.8%, 95% CI = -5.6 to -4.1) lateral ventricles and minimal change of cortical thickness in AD signature regions (left = 0.3%, 95% CI = -0.6 to 1.2; right = 0.2%, 95% CI = - 0.6 to 0.9) (Supplemental Fig. S8).

Notably, at the time of diagnosis, clinicians noted uncertainty in this diagnosis due to several complicating factors including low baseline cognitive performance with minimal decline over multiple years of evaluations, and health conditions that could contribute to cognitive impairment independently of possible AD. After the initial diagnosis, pTau217 results were at a normal level indicating that AD pathology was unlikely. Furthermore, this participant began taking a GLP-1 agonist in between her 6-month and 1-year MRI visits in our study. GLP-1 agonists reduce CSF production and intracranial pressure (Botfield et al., 2017; Krajnc et al., 2023; Mitchell et al., 2023), which may account for the reduction in ventricle size and apparent increase in regional volumes (Supplemental Fig. S8). Thus, all indications are that this is an accurate assessment of a complex, real-world instance of atypical brain aging.

Another example of atypical brain aging occurred in a cognitively unimpaired 70-74 year-old male (Figure 7). He had notably fast and asymmetric (right > left) hippocampal atrophy (left = -1.5%, 95% CI = -2.4 to -0.6; right = -5.1%, 95% CI = -5.8 to -4.4; Figures 5h and 7). This asymmetric atrophy was prominent across multiple measures (Figure 8), including the AD signature regions (Left = -0.7%, 95% CI = -1.2 to -0.1; right = -1.9%, 95% CI = -2.5 to -1.3), lateral ventricles (Left = 7.0%, 95% CI = 5.9 to 8.0; right = 9.7%, 95% CI = 8.7 to 10.7), parahippocampal cortex (Left = -1.7%, 95% CI = -3.0 to -0.5; right = -6.7%, 95% CI = -8.2 to -5.2), and temporal pole (Left = -3.4%, 95% CI = -5.6 to -1.3; right = -6.7%, 95% CI = -8.9 to -4.4). Despite accelerated atrophy, this participant was clinically assessed to be cognitively unimpaired status at follow-up. However, subjective memory complaints were noted as well as a decline in objective neuropsychological memory performance. In addition, this participant had a pTau217 value that was in the indeterminate range prior to his baseline imaging, suggesting that AD pathophysiology may be emerging.

**Figure 7.**
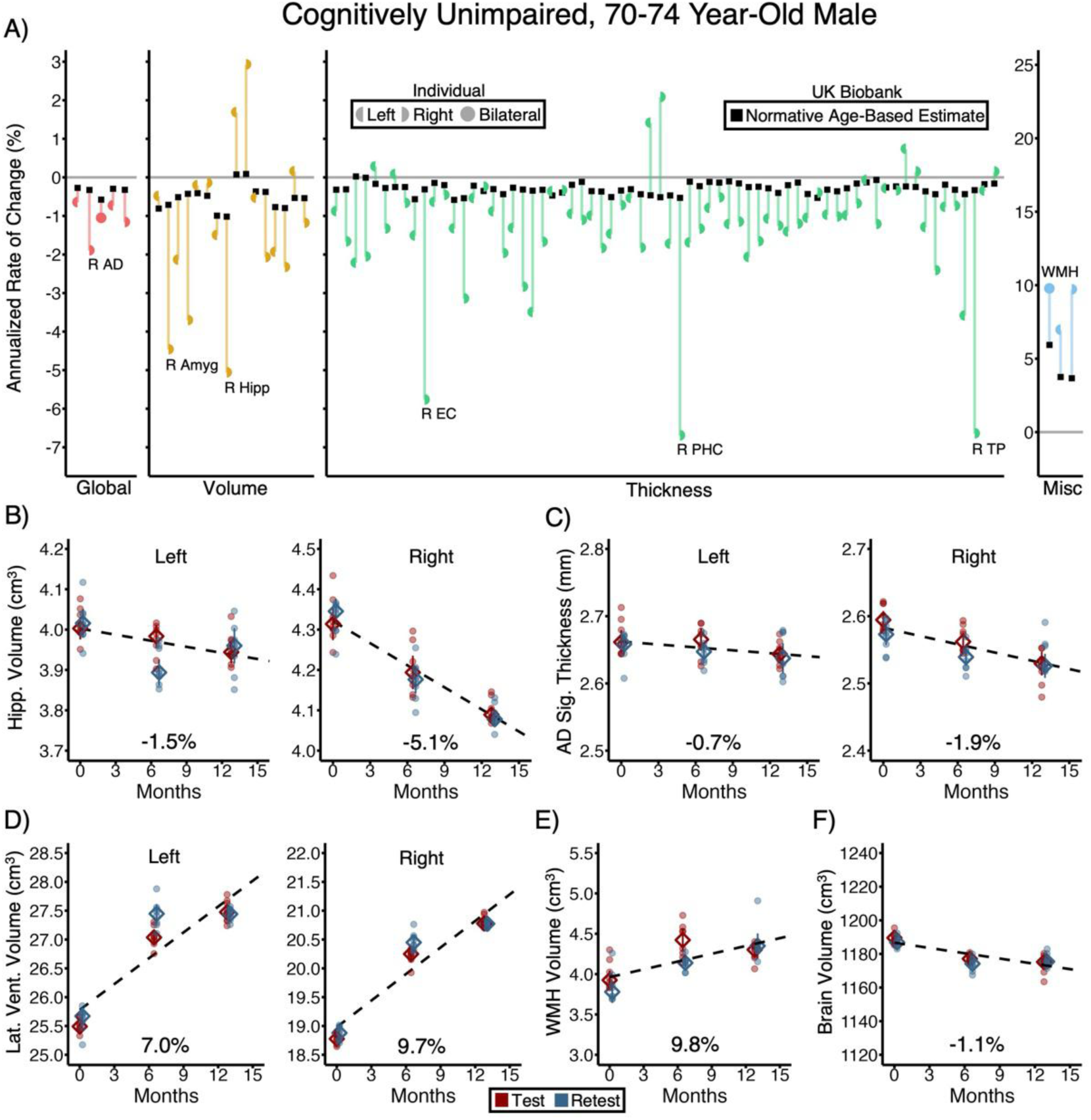
A case study of unexpected asymmetrical brain decline in a cognitively unimpaired 70-74 year-old male. Using the same format as in Figure 6, a dashboard visualization of all available brain measures for another individual showing a distinct atrophy pattern is plotted in reference to normative expectations for similarly aged individuals from the UK Biobank. All reference UK Biobank participants were within 0.8 years of this individual. The largest deviations in this individual are in multiple right-lateralized brain regions including right amygdala (Amyg) volume, right hippocampal (Hipp) volume, right entorhinal cortex (EC) thickness, right parahippocampal cortex (PHC) thickness, and right temporal pole (TP) thickness. This pattern is consistent with the earliest stages of frontotemporal lobar degeneration (FTLD). Further note that, unlike the clinically diagnosed and enrolled individual with FTLD (e.g., Figure 5d; Supplementary Fig. S7), the absolute volumes of the right and left hippocampus are similar suggesting that the accelerated longitudinal rate of change in the right hippocampus has not been occurring for an extended period before the present study. While the future progression of this individual is uncertain, the one-year longitudinal estimates captured here reveal a brain aging pattern that is atypical and robustly estimated. See Supplemental Fig. S12B for a parallel visualization that includes 95% con]idence intervals around each estimate of change.

**Figure 8.**
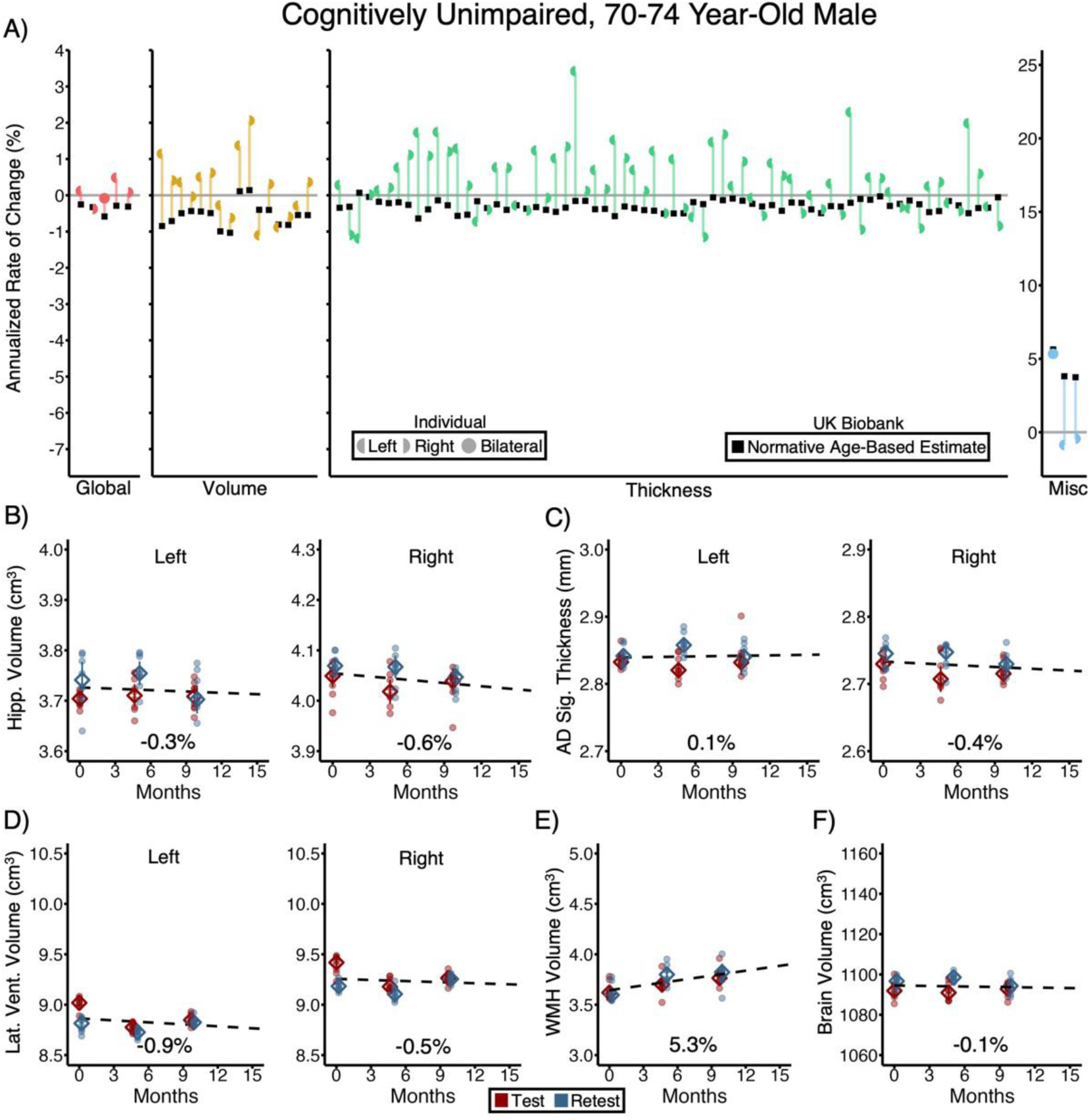
A case study of brain preservation in a cognitively unimpaired 70-74 year-old male. Using the same format as in Figures 6 and 7, a dashboard visualization of all available brain measures for another individual showing a remarkably preserved pattern of brain aging. Morphometrics are plotted referenced to normative expectations for similarly aged individuals from the UK Biobank. All reference UK Biobank participants were within 0.8 years of this individual. Note that across brain measures, this individual’s brain aging is minimal, similar to that observed for much younger adults. The detailed plots of individual measures (**B-D**) illustrate the reliability of these measures and that, across test-retest sessions, this individual is estimated to have largely preserved brain structure, atypical of individuals of his age. See Supplemental Fig. S12C for a parallel visualization that includes 95% con]idence intervals around each estimate of change.

As a final example, a cognitively unimpaired 70-74 year-old male possessed a remarkably maintained brain (Figure 8). This individual had minimal hippocampal atrophy (left = -0.3%, 95% CI = -1.2 to 0.6; right = -0.6%, 95% CI = -1.4 to 0.1; Figures 5f and 8). Brain maintenance was observed across nearly all regional volumes and cortical thickness measures, as well as maintained left (-0.9%; 95% CI = -1.9 to 0.1) and right (-0.5%; 95% CI = -1.5 to 0.5) lateral ventricles. The estimate of annualized whole brain volume decline was extremely low (- 0.1%, 95% CI = -0.4 to 0.3) and consistent across the independent data from the test and retest MRI sessions. Thus, by all accounts, this individual, living in his eighth decade, has a current brain aging trajectory roughly similar to that of a young adult and starkly different from many of his age-matched peers.

## Discussion

Precision brain aging trajectories estimated over one year revealed marked heterogeneity between people. Younger adults tended to have slower rates of brain atrophy than older adults, and older adults with MCI, AD or FTLD tended to have the fastest rates of change in brain structure. Most critically, precision estimates allowed idiosyncratic brain aging trajectories to be characterized among cognitively unimpaired individuals. Unexpected brain aging trajectories included rapid decline and asymmetric brain atrophy as well as evidence of brain maintenance. In a few extreme cases, individuals in their eighth decade showed minimal rates of atrophy similar to what one might expect in a young adult. We discuss these results in the context of moving aging research from normative, average expectations to the study of brain aging within individuals.

### Precision estimation of idiosyncratic, individualized trajectories of brain aging

The field of aging does not typically look at within-individual estimates of brain change over short intervals. Rather, brain aging is typically modeled as aggregates over many individuals or extended longitudinal intervals. Prior studies have revealed the opportunity for examining precision change within individuals. For example, in our own work, we previously collected 3-4 scans per timepoint using conventional long scans (each scan 6.6 minutes) (Fotenos et al., 2005; 2008). Over multiple years, with multiple scans, estimates of whole-brain atrophy stabilized for the largest brain morphometric measures such as whole-brain volume (see Figure 2 in Fotenos et al., 2008). Resnick and colleagues provided clear evidence of longitudinal change within individuals in the Baltimore Longitudinal Study over a follow-up period of up to a decade (mean = 5.9 years) (see Driscoll et al., 2009 Figure 1).

These promising foundations inspired us to explore approaches to measure brain change in individuals over short intervals. The advent of rapid scanning techniques that allow for dense, repeat sampling provided the key innovation to achieve precision measurement (Mair et al., 2020; Hanford et al., 2021; Elliott et al., 2023; 2024). The present findings, which used these techniques to estimate brain aging, are consistent with general expectations from group studies but also highlight idiosyncratic brain aging trajectories, including atypical patterns in unimpaired individuals, that would have been attributed to measurement error in prior work.

Specifically, we observed unimpaired individuals with rapid global decline (Figure 6), asymmetric decline (Figure 7), and notable preservation (Figure 8). These findings raise the possibility that standard longitudinal structural imaging may have underestimated the true degree of individual differences in brain aging trajectories that are present in the population and can be harnessed for predicting cognitive decline and maintenance (Di Biase et al., 2023). We demonstrate that it is possible to detect unexpected brain aging trajectories in individuals in a year. Future research with larger cohorts will be needed to describe the prevalence of these atypical brain aging trajectories, their stability over time, and their consequences for cognition. For example, there is ongoing research into the nature of brain maintenance and its role in cognitive aging and resilience to cognitive impairment (Cook et al., 2017; Katsumi et al., 2022; Kremen et al., 2022; Nilsson & Lövdén, 2018; Nyberg et al., 2012; Stern et al., 2020). Cluster scanning is a promising approach to accelerate this research.

While we found that cluster scanning provides the precision to detect differences among individuals in just one year, these results raise important questions about how stable these trajectories are over extended periods in individuals. It is unclear when in the lifespan brain aging trajectories are linear with gradual accelerations due to aging and disease, or, if studied with the right temporal sampling, brain changes may occur in fits and starts with periods of stability and rapid decline intermixed in the same individuals over time. Emerging precision research in individuals suggests that life events and exposures, including pregnancy, may impact brain development and aging trajectories (Newbold et al., 2020; Newbold & Dosenbach, 2021; Pritschet et al., 2024). To date, we have lacked the precision to describe and interrogate these dynamics in individuals. Our results provide an initial demonstration that the dynamics of brain aging and neurodegeneration can be studied over brief intervals within individuals.

Our results also raise opportunities for exploring mechanisms of cognitive maintenance and decline. In a relatively small-scale study, we captured a rapid period of brain decline in which an individual subsequently developed MCI after having been enrolled while cognitively unimpaired. This case was the most extreme case of decline in our cohort and suggests that cluster scanning can be a tool for further understanding the time course and anatomical links between brain aging trajectories and cognitive, behavioral, and emotional changes. Combining cluster scanning with emerging high-precision tools for assessing cognition and behavior in individuals, like “burst sampling”, may be fruitful (Cerino et al., 2021; Hawks et al., 2023; Papp et al., 2024; Sliwinski, 2008).

### Cluster scanning is a general strategy for stabilizing the measurement of change

Cluster scanning offers several practical advantages for longitudinal brain imaging research. With the same longitudinal design, lower measurement error will boost statistical power to enhance research into the causes and consequences of individual differences in brain development and aging (Zuo et al., 2019; Vidal-Pineiro et al., 2024). Moreover, cluster scanning can accelerate longitudinal studies because higher measurement precision allows for smaller changes that occur over shorter follow-up times to be detected. Shorter longitudinal studies can reduce participant burden, reduce costs, and limit attrition.

Rapid cluster scans each take only one minute, allowing the number of scans to be flexibly adapted to dit a study’s needs. Collecting fewer scans can minimize cost and burden, while additional rapid scans can be collected in studies where extreme precision is required. We found that estimates from eight rapid scans led to nearly a threefold reduction of longitudinal measurement error as compared to a standard scan (Elliott et al., 2024). Importantly, eight rapid scans require slightly less than 10 minutes of total scan time and thus can be incorporated into most MRI studies with minimal additional burden.

Relevant to populations who may move more in the scanner (e.g., children, individuals with brain disorders, and older adults), cluster scanning may be especially beneficial because, with multiple rapid scans, scans corrupted by head motion can be easily removed during quality control and repeated if detected while scanning. This approach can reduce sampling bias and facilitate high-quality data collection in high-value populations with a higher propensity for head motion who would often be removed from analyses with standard protocols.

## Limitations

There are several limitations. First, we found substantial heterogeneity in brain aging that was detectable due to the improved precision of cluster scanning. Future research, with clinical follow-up in larger samples, will be needed to yield translational insights tied to outcomes. Second, while the reliability of rapid estimates of brain aging trajectories was higher than standard approaches, reliability was still only moderate for many brain regions. Even higher precision is needed and could be obtained through several methods, including collecting even more rapid scans, improvements in segmentation algorithms through advances in deep learning (e.g., Heckel et al., 2024), and improved tools for deriving individualized masks of vulnerable regions (e.g., Dickerson et al., 2009). A third limitation is that our assessment of brain aging trajectories was limited to measures from T1-weighted structural MRI. Additional sensitivity may be derived from a multi-modal approach that combines measures from T2, diffusion-weighted imaging, functional MRI, and other modalities. A final limitation is that while we speculate on factors that might cause the observed heterogeneity, the present study, because of its design and scale, was unable to determine mechanisms. Cluster scanning has the potential to aid future studies that are designed to gain insight into the mechanisms of variability in brain aging.

## Conclusion

There is marked heterogeneity in brain aging across the lifespan, in brain disorders, and even among older adults of the same age. Using cluster scanning, we captured heterogeneity in individuals within one year. With this enhanced precision, surprising cases emerged, including rapid decline in cognitively unimpaired individuals and atypical maintenance. Longitudinal studies with cluster scanning promise to advance our understanding of heterogeneity in brain aging and neurodegenerative disease.

## Methods

### Estimating normative brain aging trajectories from UK Biobank participants

The UK Biobank is a prospective cohort study involving around 500,000 community volunteers recruited between 2006 and 2010 from assessment centers in England, Scotland, and Wales (Miller et al., 2016; Sudlow et al., 2015). Participants provided detailed information on demographics, lifestyle, and disease history, which was linked to electronic medical records at baseline. Since 2014, a subset has undergone brain 3T MRI scans (see Alfaro-Almagro et al., 2017; Miller et al., 2016 for MRI acquisition details), with 43,050 cross-sectional scans and an additional 4,629 longitudinal scans available at the time of our analyses (mean duration between baseline and follow-up scans = 2.63 years; range = 1.00 to 7.34 years). Participants were aged 44-82 at the time of their baseline scans.

We investigated brain morphometric measures that were derived from the T1-weighted structural scans. To ensure the inclusion of only high-quality estimates, we performed a participant selection procedure (see Supplemental Fig. S10). Participants were removed who had absolute values greater than 3 SDs from the mean of inverted signal-to-noise ratio in T1, inverted contrast-to-noise ratio in T1, volume-ratio of MaskVol-to-eTIV in the whole brain generated by subcortical volumetric segmentation, or absolute values greater than 4 SDs from the mean on eTIV, mean global cortical thickness, total surface area, scanner lateral brain position, scanner transverse brain position, scanner longitudinal brain position, and scanner table position. Individuals with a BMI higher than 40 were also removed. Finally, because our goal was to describe normative age-related variation in the population, we removed individuals who had a dementia diagnosis and individuals who carried two copies of the E4 variant of apolipoprotein E (APOE4) based on the single nucleotide polymorphism (SNP) rs429358 (chr19:44908684; T/C). After screening, the final UK Biobank dataset included 39,337 individuals in the cross-sectional sample and 4,243 in the longitudinal sample.

### Estimating precision brain aging trajectories within individuals

Fifty-eight volunteers participated in the cluster scanning study in exchange for payment. Of these, 38 completed the entire 6-visit longitudinal protocol and were included in the final sample. Adults (ages 18-49; n = 6) were recruited from the community, while older adults (ages falling between 55-89) were recruited from the Massachusetts Alzheimer’s Disease Research Center and the Frontotemporal Disorders Unit of the Massachusetts General Hospital. CDR scores (Hughes et al., 1982; Morris, 1993) and clinical diagnoses were made by experienced neurologists. Note that here and elsewhere, ages are approximated by 5-year age bins to protect confidentiality, so an age range with a maximum of 87 is approximated to have a maximum of 89 because the age falls within the 85-89 age bin, and an 87-year-old individual would be described as being 85- 89 years old. Older participants were either cognitively unimpaired (CDR = 0; n = 18) or had very mild, mild or moderate dementia (CDR = 0.5, 1 or 2; n = 14). As part of ongoing longitudinal cohorts, older participants completed annual visits to the Massachusetts Alzheimer’s Disease Research Center that were independent of our cluster scanning study. These visits included neuropsychological evaluation, informant reporting, and a formal diagnostic review by trained neurologists to determine cognitive status. Many of these participants provided single timepoint MRI data to prior analyses (e.g., Elliott et al. 2023; 2024); all longitudinal data are new and presented here for the first time.

Specific diagnoses included mild multi-domain dementia likely due to AD (probable AD validated by amyloid and tau CSF or PET biomarkers; n = 6), amnestic MCI potentially due to AD, indicated by clinical evaluation with MRI but without molecular biomarker verification (n = 3) or mild to moderate dementia due to FTLD (validated by clinical MRI and PET biomarkers; n = 5). These groups were selected to assess the feasibility of rapid scans across individuals with differing levels of age-associated atrophy, various neurodegenerative atrophy patterns, and potential movement and compliance issues of typical patient populations. All participants provided informed consent following the Mass General Brigham Healthcare Institutional Review Board guidelines. The final analyzed sample consisted of 38 participants (22 females; mean age = 64.4, range = 18-89; Table 1).

**Table 1.** Participant demographics.

| **Group** | **Sample** | **Age**<br>**Mean**<br>**(range)** | **Sex**<br>**(M/F)** | **CDR**<br>**(0/0.5/1/2)** | **CDR SOB**<br>**Mean**<br>**(range)** |
| --- | --- | --- | --- | --- | --- |
| Adults | 6 | 34.7<br>(18 - 49) | 3/3 | N/A | N/A |
| Older Adults –<br>CU | 18 | 71.6<br>(55 - 89) | 5/13 | 18/0/0/0 | 0<br>(0 - 0) |
| Older Adults –<br>MCI or AD | 9 | 68.9<br>(55 - 79) | 5/4 | 0/4/5/0 | 4.78<br>(0.5 - 9) |
| Older Adults –<br>FTLD | 5 | 65.8<br>(50 - 79) | 3/2 | 1/2/1/1 | 3.40<br>(0.5 - 8) |

Notes: CDR = clinical dementia rating; SOB = sum of boxes; M = male; F = female; CU = cognitively unimpaired older adult; MCI = mild cognitive impairment; AD = Alzheimer's disease; FTLD = frontotemporal lobar dementia. The age range is approximate to reflect 5-year bins (e.g., a maximum age of 87 would lead to a maximum age range of 89 because the bin is 85-89).

### Plasma collection and analysis

As part of their annual visits at the Massachusetts Alzheimer’s Disease Research Center, plasma samples were collected and processed for biomarker analysis in a subset of the older adult participants. Of the 18 CDR 0 older participants who were recruited from the Massachusetts Alzheimer’s Disease Research Center, 11 had a plasma sample that was collected within 5 years of their baseline MRI scan in our study. Of the 9 older participants with MCI or Alzheimer’s Dementia who were recruited from the Massachusetts Alzheimer’s Disease Research Center, 6 had a plasma sample that was collected within 5 years of their baseline MRI scan in our study. These plasma samples were assayed for pTau217 to determine the presence of Alzheimer’s Disease pathology.

Plasma samples were collected in K2EDTA tubes, centrifuged and frozen within 4 hours of collection, and stored at -80°C until use. pTau217 was measured in duplicate without dilution using the MSD S-PLEX pTau217 (cat#: K151APFS) assay at an MSD laboratory following the manufacturer’s instructions. Samples were part of a larger study and were randomly distributed over 93 plates. Three internal control samples spanning the assay range were included in duplicate on each plate to account for plate-to-plate variability. The standardized mean of the internal control samples all fell within the prespecified acceptance limit of 80-120% of the mean of all controls and no normalization was considered necessary. Intraplate replicate CVs were 8.0±11.0% (mean±SD) and interplate CVs for the internal control samples were 8.0±0.9%.

Thresholds to differentiate individuals with and without AD pathology were established in-house using 320 participants from the Massachusetts Alzheimer’s Disease Research Center longitudinal cohort study with molecular biomarker-verified AD status (PET, CSF, and/or postmortem histopathological examination). This established a binary threshold for AD of 6.1 pg/ml using the Youden index (AUC 0.94) with an indeterminate range 4.95-8.97 pg/ml (92.5% sensitivity and specificity).

### MRI data acquisition for cluster scanning

MRI data were acquired using a 3T Siemens MAGNETOM Prisma^*it^ MRI scanner (Siemens Healthineers AG; Erlangen, Germany) at the Harvard University Center for Brain Science, equipped with the vendor’s 32-channel head coil. The scanner and the Alzheimer’s Disease Neuroimaging Initiative (ADNI) protocols were validated under the Standardized Centralized Alzheimer’s & Related Dementias Neuroimaging Initiative (https://scan.naccdata.org/). During the scans, participants could watch video clips, such as nature documentaries, or listen to music. To enhance hearing protection and stabilize the participants’ heads, inflatable cushions were employed. Reminders to stay still and feedback on head motion were provided to participants every 5-10 minutes.

Two different T1-weighted acquisitions were collected: (1) 1.0 mm isotropic ADNI3 MPRAGE acquisition (5’12’’ acquisition; pulse repetition time (TR) = 2300 ms; inversion time (TI) = 900 ms; time to echo (TE) = 2.98 ms; flip angle = 9°; field of view = 256 x 240 x 208 mm; acquisition orientation = sagittal; in-plane GRAPPA acceleration = 2)(Weiner et al., 2017), (2) 1.0 mm isotropic compressed sensing scans (1’12’’ acquisition; TR= 2300 ms; TI = 900 ms; TE = 2.96 ms; flip angle = 9°; field of view = 256 × 192 × 240 mm; acquisition orientation = coronal; compressed sensing acceleration = 6x)(Mussard et al., 2020). We refer to these scans as the “standard” and “rapid” scans, respectively. Turbo Factor/Samples-per-TR in the rapid sequence was kept approximately constant at a value close to that used in the standard scan to avoid effects from differential T1 weighting during the readout train. A coronal acquisition was employed for the rapid scans, in contrast to the sagittal acquisitions in the standard scan, as sagittal acquisition orientation compounded susceptibility-induced artifacts in the orbitofrontal cortex (Hanford et al., 2021).

### Separation of longitudinal change from measurement error

The study was designed to measure the morphometric brain changes that occur in approximately one year and to simultaneously assess measurement error. Each participant completed six MRI sessions spread over three timepoints. In the first half of each scanning session, a single standard scan was collected alongside a “cluster scanning” block of four rapid scans. Then, each participant was briefly taken out of the scanner to stretch and use the restroom. After the break, the participant was repositioned, and the scanner re-shimmed. An identical set of four rapid scans was repeated, yielding a total of one standard scan and eight rapid scans collected in each scan session, and a total of 6 standard scans and 48 rapid scans across the longitudinal study (Figure 1). The order of the standard and rapid scans was counterbalanced between participants to mitigate order effects.

Critically, to assess test-retest measurement error, participants completed a pair of scanning sessions on closely spaced separate days at baseline, as well as at approximately six months and one year later. The 6 visits were completed across a mean of 11.6 months (range = 7.7 to 14.3 months). The mean interval between test and retest sessions was 7.8 days (range = 1 to 23 days). This sampling design allowed us to directly measure the test-retest error of longitudinal change because two independent estimates of the rate of change could be calculated (from sessions 1, 3, and 5 and separately from sessions 2, 4, and 6; see Figure 1). To our knowledge, this is the first longitudinal MRI study to implement this design, which allows explicit measurement of the longitudinal test-retest error from independent but temporally aligned data samples collected within each individual.

### Image processing and morphometry

All structural images from both the UK Biobank and the cluster scanning study were analyzed using FreeSurfer version 6.0.1 and the recon-all processing pipeline (Dale et al., 1999; Fischl et al., 1999). Each scan was processed through the cross-sectional recon-all pipeline. All longitudinal data were also processed through the FreeSurfer longitudinal pipelines. Quality control was conducted by visually inspecting all structural images to note motion artifacts, banding, ringing, and blurring. All scans from the 38 participants with longitudinal data passed quality control criteria and were included in all analyses.

#### Cross-sectional pipeline

In the volume-based processing, intensity normalization, skull stripping, and segmentation of regional brain volumes were performed (Fischl et al., 2002; Ségonne et al., 2004). Subsequently, models of the white-matter surface and pial surface were created from each scan using the surface-based processing pipeline (Dale et al., 1999; Fischl et al., 1999). The results from the cross-sectional pipeline were used to examine cross-sectional brain differences in the data from the UK Biobank as well as estimate the nominal test-retest error of the measurements in the cluster-scanning data from each timepoint (where the data from the two separate days were analyzed independently).

#### Longitudinal pipeline

To estimate the rates of longitudinal brain change, scans from participants with more than one timepoint were separately processed through FreeSurfer’s longitudinal recon-all processing pipeline, which consists of two primary steps (Reuter et al., 2012). First, unbiased within-subject templates were created for each participant (Reuter & Fischl, 2011). Second, several recon-all processing steps were recomputed using the within-individual template to initialize the processing (Reuter et al., 2012). We generated several longitudinal templates for each participant so that we could derive fully independent and unbiased test and retest estimates of morphometric measures for analyses of longitudinal measurement error. Specifically, from the three test sessions (sessions 1, 3, 5) we generated the following five subject-specific individualized templates from the following scans from each participant: (1) one standard scan from each test session (a total of three standard scans per participant), (2) one rapid scan from each test session (a total of 3 rapid scans from each participant), (3) two rapid scans from each test session (a total of 6 rapid scans from each participant), (4) four rapid scans from each test session (a total of 12 rapid scans from each participant), and (5) eight rapid scans from each test session (a total of 24 rapid scans from each participant). In parallel, we also generated the same five templates from the retest sessions (sessions 2, 4, and 6). Finally, we also generated a template from all eight rapid scans that were collected across each of the six sessions (a total of 48 rapid scans from each participant) to investigate the highest precision cluster scanning measurements in our study. This resulted in a total of 11 templates (five from test sessions, five from retest sessions, and one combining all test and retest sessions) for each participant. Morphometric measures for the included scans were then extracted separately for each template.

Importantly, no estimates in either the cross-sectional or longitudinal processing pipelines were manually adjusted, allowing the automated metrics to deliver an unbiased assessment of measurement error.

### Longitudinal change and measurement error

Measurement error was estimated for 82 separate brain morphometric measures. These included 14 subcortical volumes (left and right estimates of the amygdala, pallidum/globus pallidus, caudate nucleus, hippocampus, putamen, thalamus, and ventral diencephalon volume from the Aseg atlas; Fischl et al., 2002) and 68 regional cortical thickness measures (all cortical regions from the Desikan-Killiany atlas; Dale et al., 1999; Desikan et al., 2006). We used these 82 regional morphometric measures to generate summaries of error and error reduction afforded by pooling of rapid scans. In addition, we utilized the following metrics from the Aseg atlas: total brain volume (BrainSegVolNotVentSurf), left and right lateral ventricle volume and white matter hypointensities. We also measured the average cortical thickness in the AD signature (Dickerson et al., 2009).

For each scan type and morphometric measure, we first estimated the measurement error from outputs of the cross-sectional FreeSurfer pipeline. Measurement error was calculated as the absolute difference between each measure estimated from Sessions 1 versus 2, Sessions 3 versus 4, and Sessions 5 versus 6. All session pairs were collected on different days near to one another, such that minimal true change was expected, and differences in the measures would be due to noise. Larger errors indicate a greater difference between morphometric measures from each session and a higher proportion of the measurement that is attributable to error (i.e., lower precision).

In all participants, individualized linear regression models were then used to estimate the rate of change in each morphometric estimate, extracted from the longitudinal FreeSurfer pipeline. Rate of change estimates were generated by modeling change as a function of the number of years since the baseline at each MRI observation. This model yielded a regression coefficient in units of annualized rate of brain change. In analyses that contrasted measures, this annual rate was expressed as a percent change per year by dividing the regression coefficient by the mean value of the brain morphometric measure across all observations in the respective analysis.

To aid interpretation, test-retest reliability was estimated for both standard and cluster scanning protocols using the Pearson correlation for both cross-sectional and longitudinal change estimates (See Figure 3 and supplemental Figures S3-S6). Each test-retest reliability calculation was also performed using the intraclass correlation coefficient (ICC) to confirm convergent results between statistics. Specifically, ICC(3,1) was used, and nearly identical results were obtained (all ICC values within +/- 0.02 of Pearson values).

A key challenge with individualized analyses is accurately determining confidence in the individualized estimates of change. Apparent individual differences in brain aging trajectories could be due to true heterogeneity or noisy estimates. We addressed this challenge through our longitudinal test-retest design. First, in Figure 4 and throughout our results, we quantified the precision in each participant’s estimates with 95% confidence intervals from our individualized regression estimates of change. Second, we visualized the precision of our estimates by showing individual scan estimates that went into each individualized regression model (see Figures 5, 6, 7, and 8). Estimates from the test (red) and retest (blue) visits were plotted separately to transparently display the stability over independent days and the dispersion among individual scans on a given day.

### Estimating normative reference values tailored to each individual

A challenge in assessing brain aging is that brain measures show non-linear rates of change across the lifespan, and the exact rates of change differ across measures (e.g., Fjell et al., 2013; Bethlehem et al., 2022). To contextualize the individualized estimates of the rate of change, we compared individualized precision estimates from the cluster scans in the 38 intensively sampled individuals to normative longitudinal change expectations from the 4,243 UK Biobank participants.

For each participant, an individualized expectation of change was generated for every morphometric measure using 300 individuals that were sub-selected from the larger sample of 4,243 individuals. We quantified the normative estimate for each morphometric as the mean rate of change in that brain morphometric measure in the 300 UK Biobank participants closest in age to each participant. This individualized normative expectation allowed us to compare each participant’s individualized estimate of change across a year to the expected rate of change in same-aged individuals from the UK Biobank and to do so comprehensively for every measure, providing a detailed, contextualized dashboard view of each individual’s idiosyncratic pattern of brain aging (see Figures 6, 7, and 8).

## Data Availability

Novel MRI data used in this manuscript and the analysis code will be made available after publication. A description of the procedure to access the UK Biobank data is available at https://www.ukbiobank.ac.uk/enable-your-research.

## Acknowledgments

We thank the participants in this research, without whose effort it would not have been possible to accomplish this study. The UK Biobank analyses were conducted under application number 67237. This research was supported by NIA grants R01AG067420, R01DC014296, and P30AG062421, NIH Shared Instrumentation Grant S10OD020039, the Simons Foundation grant 811255, and the Vranos Family Fund. M.L.E. is supported by NIA grant K00AG068432. We thank the Harvard Center for Brain Science neuroimaging facility and the Martinos Center Compute Cluster. We thank Kayla Ntoh, Stephanie Kaiser, Joanna Ladoupolou, Francesca Davy-Falconi, and Rachel Lemley for assisting in data collection. We thank Timothy O’Keefe for assisting in data management. We thank Tom Hilbert, Tobias Kober, and the Siemens Healthineers for providing compressed sensing capabilities under a research application sequence research agreement. We thank the personnel of the Massachusetts Alzheimer’s Disease Research Center and the Frontotemporal Disorders Unit for their assistance in recruitment.

## Ethics Statement

The study protocol was approved by the Institutional Review Board of Mass General Brigham Healthcare. All participants provided written informed consent in accordance with the guidelines of the Institutional Review Board of Mass General Brigham Healthcare and were compensated. The UK Biobank received ethical approval from the Northwest Centre Research Ethics Committee (REC number 11/NW/0382).

## Credit Authorship Contribution Statement

**Maxwell L. Elliott:** Conceptualization, Methodology, Software, Investigation, Data curation, Writing – original draft, Visualization, Funding acquisition. **Jingnan Du:** Methodology, Software, Writing – review & editing, Visualization. **Jared A. Nielsen:** Methodology, Software, Writing – review & editing, Visualization. **Lindsay C. Hanford:** Methodology, Software, Writing – review & editing, Visualization. **Pia Kivisäkk:** Resources, Writing – review & editing. **Steven E. Arnold:** Resources, Writing – review & editing. **Bradford C. Dickerson:** Resources, Writing – review & editing. **Ross W. Mair:** Resources, Methodology, Data curation, Writing – review & editing. **Mark C. Eldaief:** Resources, Writing –review& editing. **Randy L. Buckner:** Conceptualization, Methodology, Resources, Data curation, Writing – review & editing, Supervision, Project administration, Funding acquisition.

